# Influence of Prior Probability Information on Large Language Model Performance in Radiological Diagnosis

**DOI:** 10.1101/2024.08.27.24312693

**Authors:** Takahiro Fukushima, Ryo Kurokawa, Akifumi Hagiwara, Yuki Sonoda, Yusuke Asari, Mariko Kurokawa, Jun Kanzawa, Wataru Gonoi, Osamu Abe

**Affiliations:** Department of Radiology, Graduate School of Medicine, The University of Tokyo, 7-3-1 Hongo, Bunkyo-ku, Tokyo, 113-8655, Japan

## Abstract

**Background:** Large language models (LLMs) show promise in radiological diagnosis, but their performance may be affected by the context of the cases presented.

**Purpose:** To investigate how providing information about prior probabilities influences the diagnostic performance of an LLM in radiological quiz cases.

**Materials and Methods:** We analyzed 322 consecutive cases from Radiology’s “Diagnosis Please” quiz using Claude 3.5 Sonnet under three conditions: without context (Condition 1), informed as quiz cases (Condition 2), and presented as primary care cases (Condition 3). Diagnostic accuracy was compared using McNemar’s test.

**Results:** The overall accuracy rate significantly improved in Condition 2 compared to Condition 1 (70.2% vs. 64.9%, p=0.029). Conversely, the accuracy rate significantly decreased in Condition 3 compared to Condition 1 (59.9% vs. 70.2%, p<0.001).

**Conclusion:** Providing context about prior probabilities significantly affects the diagnostic performance of the LLM in radiological cases. This suggests that LLMs may incorporate Bayesian-like principles in their diagnostic approach, highlighting the potential for optimizing LLM’s performance in clinical settings by providing relevant contextual information.

**Key Results:** LLM’s overall accuracy improved from 64.9% to 70.2% when informed about quiz case nature (p=0.029).

LLM’s overall accuracy decreased to 59.9% when presented with incorrect primary care context (p<0.001).

Results suggest LLMs may utilize Bayesian-like principles in diagnostic reasoning, similar to human radiologists.

**Summary Statement:** Providing context about prior probabilities significantly influences LLM’s diagnostic performance in radiological cases, suggesting potential for optimizing LLM use in clinical practice through contextual information.

## Introduction

Large language models (LLMs) are neural network models trained on huge amounts of text data and have shown excellent performance in natural language processing tasks. They are used in a variety of fields, including medicine, and attempts to utilize LLMs in radiological diagnosis have begun as follows.

In a study by Ueda et al., GPT-4 (OpenAI, San Francisco, USA) achieved over 50% accuracy rate, correctly answering 170 out of 313 cases of various disease categories in Radiology’s “Diagnosis Please” quiz cases based on the patient’s history and the imaging findings (1). Similarly, Horiuchi et al. reported that the GPT-4 achieved a diagnostic accuracy of 50% (50/100 cases) in the American Journal of Neuroradiology’s “Case of the Week” quiz series of neuroradiology cases (2). Furthermore, recent studies have demonstrated a significant improvement in the diagnostic capabilities of LLMs when provided with key images in addition to textual information (3). Comparisons have been made both within and between vendors (4,5), as well as between LLMs and human radiologists (6). While current LLMs do not yet match the diagnostic accuracy of human radiologists, they have shown potential as powerful supportive tools.

To enhance LLM diagnostic performance and facilitate future clinical applications, it is crucial to accurately evaluate their capabilities and understand their characteristics. Previous studies have utilized case collections well-suited for research, such as “Diagnosis Please” cases in Radiology, which provide comprehensive clinical histories, radiological images, and confirmed final diagnoses (logically deducible).

However, it is important to emphasize that the above studies have consistently placed LLMs at a disadvantage by withholding information about prior probabilities—an unwritten rule that human radiologists inherently consider. Quiz cases and real clinical scenarios differ substantially in terms of prior probabilities, which are vital contributors to diagnosis. Human radiologists understand that expert-level quiz cases are more likely to feature rare diseases, uncommon presentations of common diseases, educational mimickers, or recently discovered conditions, rather than typical presentations of common diseases like cerebral infarction or subarachnoid hemorrhage. In real-world clinical settings, disease prevalence varies between primary care clinics and tertiary referral hospitals (7), and human radiologists can adjust their diagnostic approach based on institutional and regional characteristics. Bayes’ theorem, which represents the probabilistic nature of clinical reasoning (8), demonstrates that accurate recognition of current circumstances affecting prior probabilities is essential for improving diagnostic accuracy, as these are crucial determinants of posterior probabilities.

Given this context, we hypothesized that providing LLMs with additional information that influences the prior probabilities of diseases in the target patient group/cohort would affect their diagnostic performance. Furthermore, we posited that providing accurate versus inaccurate additional information would lead to variations in diagnostic performance for the same set of cases. The aim of this study is to prove these hypotheses and further elucidate the characteristics of LLMs in medical diagnosis.

## Materials and Methods

Ethical approval was not required as this study exclusively used data from previously published articles.

We used clinical history and author-provided figure legends of 322 consecutive cases (from Aug 1 1998 to Oct 31 2023) from Diagnosis Please, a monthly quiz case collection for diagnostic imaging physicians published in Radiology.

We used Claude 3.5 Sonnet (Anthropic, San Francisco, United States; released on June 27, 2024) to list the primary diagnoses and two differential diagnoses for the cases. Application programming interfaces were used to access the model (Claude 3.5 Sonnet: claude-3-5-sonnet-20240620) on August 17, 2024. To ensure reproducibility, we specified the generation parameters for all models as temperature□=□0.0. To prevent previous inputs from influencing subsequent ones, inputs were conducted in an independent session for each case for each condition.

We used three different prompts as follows:

Condition 1: not given as being a quiz case and no situation is set: “Assuming you are a physician, please respond with the most likely diagnosis and the next two most likely differential diagnoses based on the attached information.”

Condition 2: given as being quiz cases: The prompt for Condition 1 with “This is a quiz case for diagnostic radiologists, and your goal is to correctly answer this quiz case. In this quiz case, diseases with concepts established within the last 5 years, rare diseases, rare presentations of common diseases, and educational mimickers are more likely to be asked, while typical presentations of common diseases are less likely to be questioned.” Condition 3: presented as a primary care situation: “You are an experienced primary care physician. You are examining a patient who has come to your primary care clinic. Please respond with the most likely diagnosis and the next two most likely differential diagnoses based on the attached information.”

Each prompt was submitted to the model only once, and the first response generated was used for evaluation. The accuracy of the primary diagnosis and two differential diagnoses generated by the models were determined by consensus between one trainee radiologist and one board-certified diagnostic radiologist with 11 years of experience. McNemar’s test was used to assess the difference in correct response rates for the overall accuracy between Conditions 1, 2, and 3. Two-sided p-values□< □0.05 were considered statistically significant. Statistical analyses were performed using R (version 4.1.1; R Foundation for Statistical Computing, Vienna, Austria).

## Results

The overall accuracy rate was significantly improved in Condition 2 compared with Condition 1 (226/322 (70.2%) vs. 209/322 (64.9%), p = 0.029). Conversely, the overall accuracy rate was significantly decreased in Condition 3 compared with Condition 1 (193/322 (59.9%) vs. 226/322 (70.2%), p <0.001) (**Table 1**).

**Table 1.**
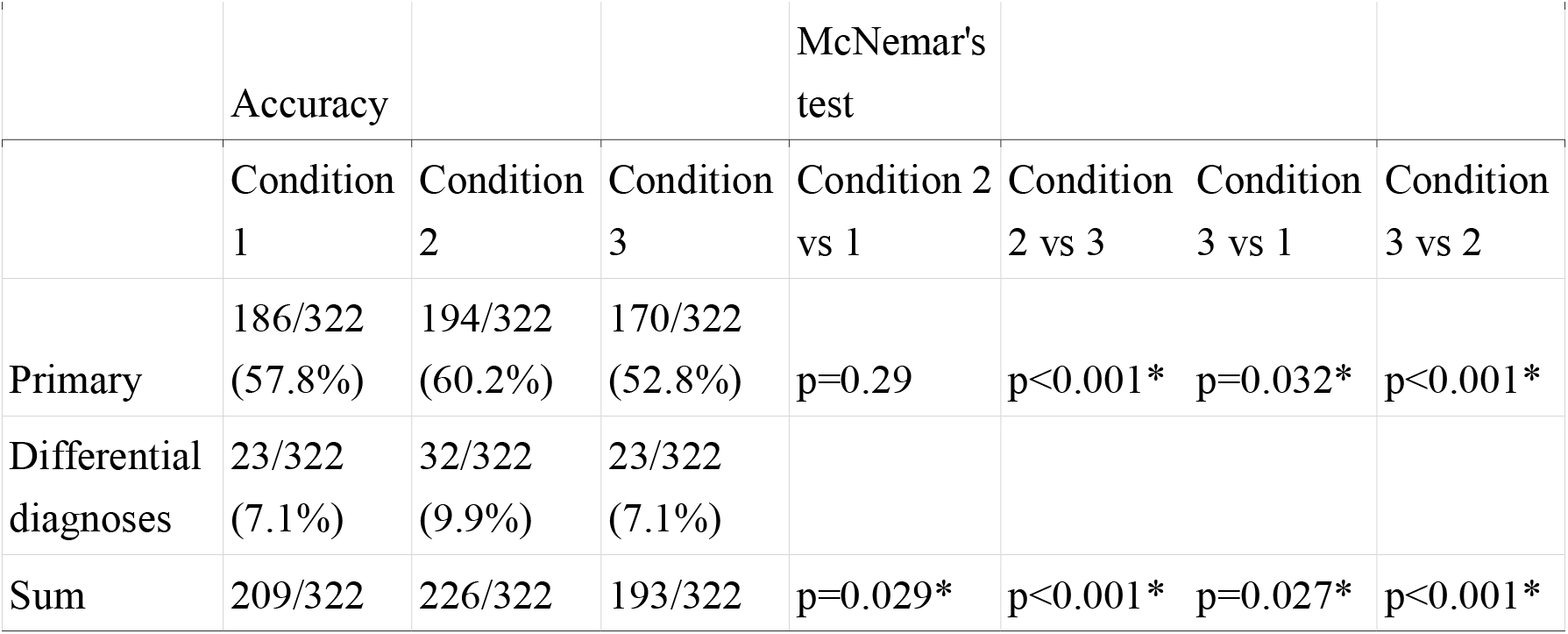
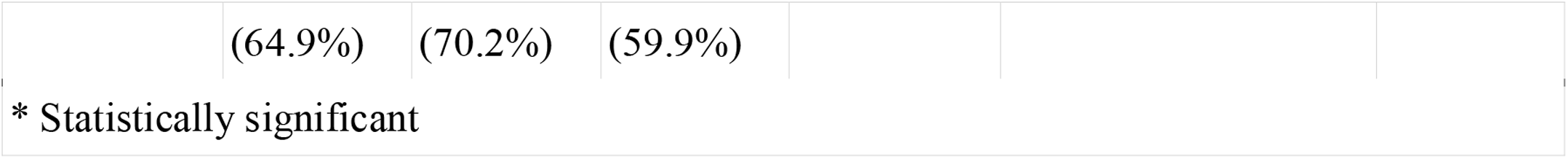
Results.

|  | Accuracy |  |  | McNemar's test |  |  |  |
| --- | --- | --- | --- | --- | --- | --- | --- |
|  | Condition 1 | Condition 2 | Condition 3 | Condition 2 vs 1 | Condition 2 vs 3 | Condition 3 vs 1 | Condition 3 vs 2 |
| Primary | 186/322 (57.8%) | 194/322 (60.2%) | 170/322 (52.8%) | $p=0.29$ | $p<0.001^*$ | $p=0.032^*$ | $p<0.001^*$ |
| Differential diagnoses | 23/322 (7.1%) | 32/322 (9.9%) | 23/322 (7.1%) |  |  |  |  |
| Sum | 209/322 | 226/322 | 193/322 | $p=0.029^*$ | $p<0.001^*$ | $p=0.027^*$ | $p<0.001^*$ |

|  |  |  |  |
| --- | --- | --- | --- |
|  | (64.9%) | (70.2%) | (59.9%) |
| * Statistically significant |  |  |  |

Examples where listing common diseases in Condition 1 resulted in incorrect answers, but listing rare diseases in Condition 2 led to correct answers are summarized in **Table 2** (9,10).

**Table 2.** Examples where differences between correct and incorrect responses under Conditions 1 and 2.

|  | Correct answer | Primary diagnosis under Condition 1 (incorrect answer) | Differential diagnoses under Condition 1 (incorrect answer) | Correct diagnosis under Condition 2 |
| --- | --- | --- | --- | --- |
| Case 166 | Metastatic Left Pulmonary Artery Sarcoma | Metastatic cancer | Primary lung cancer, Pulmonary embolism | Pulmonary artery sarcoma |
| Case 203 | Gorham Disease | Intraosseous Hemangioma | Multiple Myeloma, Metastatic Carcinoma | Gorham-Stout disease |

Examples where listing uncommon diseases in Condition 1 resulted in correct answers, but listing common diseases in Condition 3 led to incorrect answers are summarized in **Table 3** (11).

**Table 3.**
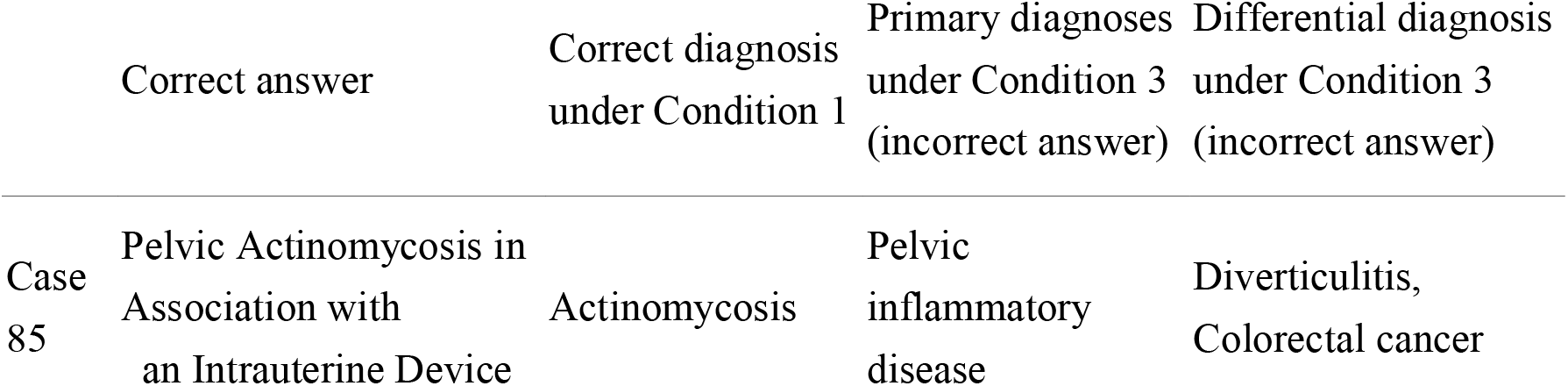
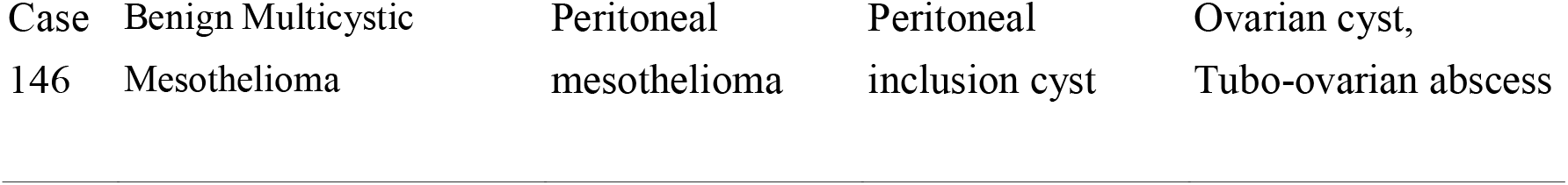
Examples where differences between correct and incorrect responses arose under Conditions 1 and 3.

## Disccusion

In this study, we utilized cases from Radiology’s “Diagnosis Please,” a radiological image diagnosis quiz for radiologists. When provided with additional information influencing prior probabilities, specifically informing Claude 3.5 Sonnet that these were quiz cases, we observed a significant improvement in the diagnostic performance of the LLM (overall accuracy rate increased from 64.9% to 70.2%). Conversely, when given incorrect additional information, such as setting the context as a primary care clinic, a significant decrease in diagnostic performance was observed (overall accuracy rate decreased from 70.2% to 59.9%).

Accurate performance evaluation and understanding of LLM characteristics are crucial for exploring their potential applications. Case collections like “Diagnosis Please,” which provide detailed clinical histories, radiological images, and confirmed final diagnoses that can be logically deduced from the given information, are well-suited for LLM performance evaluation (e.g., comparisons between vendors or versions within the same vendor) and assessing similarities and differences with human radiologists. Various authors have previously conducted such studies (1,3,4,6,12,13). However, while previous studies have assigned the role of a radiologist to LLMs, they have not informed the LLM that the cases were from quizzes. We believe this created a non-negligible gap that should be addressed.

Human radiologists can judge which answers are more likely to be correct based on the situation. For example, they can appropriately adjust the gradient of prior probabilities based on premises such as the type of medical facility (primary care vs. tertiary hospital), region, or whether it is a quiz case, thereby improving their diagnostic accuracy (8,14). The importance of prior probabilities in determining the final diagnosis is supported by Bayes’ theorem (8). Bayes’ theorem represents the probabilistic nature of diagnostic reasoning in a mathematical formula and is generally expressed in the context of clinical reasoning using posterior probabilities (predictive values), sensitivity (Sn) and specificity (Sp), and prior probabilities (often called prevalence; Prev) as follows (8):

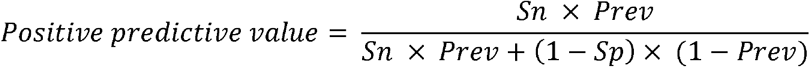

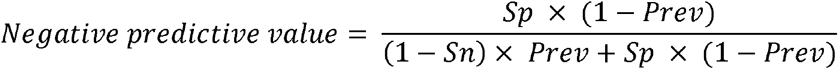

This equation represents the relationship between probabilities acting in the diagnostic process, demonstrating that the critical determinants of posterior probabilities for a disease are the prior probabilities and test characteristics. The Bayesian approach has been known to be useful for diagnosing rare diseases; as new evidence emerges, clinicians can update their prior probabilities and identify rare diseases that initially seemed unlikely (15).

We hypothesized that Bayes’ theorem might play a crucial role not only in human diagnosis but also in the diagnostic process of state-of-the-art LLMs. Specifically, we predicted that providing information as prompts to assist in estimating prior probabilities based on this theorem would enhance the diagnostic performance of Claude 3.5 Sonnet. Conversely, we anticipated that setting a context where encountering cases likely to be the correct answers in quizzes is infrequent would decrease diagnostic performance. The results supported our hypothesis. In Condition 2, where we provided prompts that aligned the assumed prior probabilities with the situation, emphasizing rare diseases, the performance improved. In contrast, in Condition 3, where we provided prompts that deviated from this assumption by emphasizing common diseases, the performance declined. Although the reasoning process of LLMs remains a black box, these results suggest that in the context of clinical reasoning, providing prompts that influence prior probabilities produces outcomes analogous to the effects on human clinical reasoning. This finding implies that LLMs may be incorporating Bayesian-like principles in their diagnostic approach, even though the exact mechanisms remain unclear.

The results of this study suggest several directions for future research. Just as informing the LLM about the quiz nature of the cases improved its diagnostic performance, providing context about real clinical situations may enhance LLM performance in actual clinical settings. For instance, supplying the LLM with information from databases on prevalence rates specific to regions or institutions could optimize its diagnostic results for those particular settings, making it more valuable in our practice. This underscores the growing importance of developing databases for individual regions and institutions. Proper ethical review processes will be essential to enable the input of clinical data into LLMs.

This study has several limitations. The analysis was based on a limited number of cases, precluding subgroup analysis by disease categories. As the answer criteria set by the “Diagnosis Please” creators are not publicly available, our judgments of correct and incorrect answers may differ from the actual standards. Additionally, since all cases have been published as papers, there is a possibility that they were used in training the LLM.

## Conclusion

We demonstrated that in radiological image diagnosis quizzes, providing prior information about the quiz nature of the cases significantly improved the diagnostic performance of Claude 3.5 Sonnet. Conversely, giving incorrect context, such as a primary care setting, significantly decreased its performance. Similar to human physicians, the concept of prior probability, as suggested by Bayes’ theorem, appears to be crucial for the LLM. This implies that constructing and providing optimized databases for specific regions and institutions to LLMs could enhance their diagnostic performances, potentially allowing LLMs to contribute more substantially to real clinical practice.

## Data Availability

This study exclusively used data from previously published articles.

## Abbreviations

LLM: large language model

## References

1. Ueda D, Mitsuyama Y, Takita H, et al. ChatGPT’s Diagnostic Performance from Patient History and Imaging Findings on the Diagnosis Please Quizzes. Radiology. 2023;308(1):e231040.

2. Kihara M, Ikeda Y, Shibata K, et al. Pharmacokinetic profiles of intravenous imipenem/cilastatin during slow hemodialysis in critically ill patients. Clin Nephrol. 1994;42(3):193–197.

3. Kurokawa R, Ohizumi Y, Kanzawa J, et al. Diagnostic performances of Claude 3 Opus and Claude 3.5 Sonnet from patient history and key images in Radiology’s “Diagnosis Please” cases. Jpn J Radiol. 2024; doi: 10.1007/s11604-024-01634-z.

4. Sonoda Y, Kurokawa R, Nakamura Y, et al. Diagnostic performances of GPT-4o, Claude 3 Opus, and Gemini 1.5 Pro in “Diagnosis Please” cases. Jpn J Radiol. 2024; doi: 10.1007/s11604-024-01619-y.

5. Cuccagna C, Schinzari F, Piccininni C, et al. Post-surgical spontaneous paroxysmal hypothermia: a case series. Clin Auton Res. 2024; doi: 10.1007/s10286-024-01048-x.

6. Horiuchi D, Tatekawa H, Oura T, et al. Comparing the Diagnostic Performance of GPT-4-based ChatGPT, GPT-4V-based ChatGPT, and Radiologists in Challenging Neuroradiology Cases. Clin Neuroradiol. 2024; doi: 10.1007/s00062-024-01426-y.

7. Uy EJB. Key concepts in clinical epidemiology: Estimating pre-test probability. J Clin Epidemiol. 2022;144:198–202.

8. Bours MJ. Bayes’ rule in diagnosis. J Clin Epidemiol. 2021;131:158–160.

9. Mittra ES, Iagaru AH, Leung AN. Case 166: Metastatic left pulmonary artery sarcoma. Radiology. 2011;258(2):645–648.

10. de Villiers JFK, Stevens WR. Case 203: Gorham disease. Radiology. 2014;270(3):931–935.

11. Lely RJ, van Es HW. Case 85: pelvic actinomycosis in association with an intrauterine device. Radiology. 2005;236(2):492–494.

12. Horiuchi D, Tatekawa H, Shimono T, et al. Accuracy of ChatGPT generated diagnosis from patient’s medical history and imaging findings in neuroradiology cases. Neuroradiology. 2024;66(1):73–79.

13. Oura T, Tatekawa H, Horiuchi D, et al. Diagnostic accuracy of vision-language models on Japanese diagnostic radiology, nuclear medicine, and interventional radiology specialty board examinations. Jpn J Radiol. 2024; doi: 10.1007/s11604-024-01633-0.

14. Westbury CF. Bayes’ rule for clinicians: an introduction. Front Psychol. 2010;1:192.

15. Gill CJ, Sabin L, Schmid CH. Why clinicians are natural bayesians. BMJ. 2005;330(7499):1080–1083.

